# Determinants of COVID-19 data quality in the District Health Information Management System version 2 in Ahafo Region, Ghana

**DOI:** 10.1101/2024.05.08.24307079

**Authors:** Felix Gumaayiri Aabebe, Silas Adjei-Gyamfi, Clotilda Asobuno, Samuel Malogae Badieko, Blaise Bagyliku Kuubabongnaa, Samuel Kwabena Boakye-Boateng, Tsunoneri Aoki, Hirotsugu Aiga

## Abstract

**Background:** In ensuring public health efforts in combating pandemics such as coronavirus disease 2019 (COVID-19), transparent data reporting that is of high quality and easily accessible is crucial for tracking epidemic progress and making informed decisions. In Ghana, no published studies have been conducted to evaluate the quality of COVID-19 data submitted onto the national web-based platform, District Health Information Management System version 2 (DHIMS-2) during the COVID-19 period. In this regard, this study seeks to assess the estimates and determinants of COVID-19 data quality in the DHIMS-2 in the Ahafo region of Ghana.

**Methods:** A facility-based cross-sectional study design was employed, with a desk review of COVID-19 records in DHIMS-2 and primary data sources (registers and monthly reporting forms). This study involved all 23 different levels of healthcare facilities that reported on COVID-19 in the Ahafo region from March 2020 to December 2022. We assessed COVID-19 data quality using three dimensions of completeness, accuracy, and timeliness according to the World Health Organization data quality guide. Mixed-effect logistic regression was then employed to identify the determinants of COVID-19 data quality at a 95% confidence interval.

**Results:** The overall COVID-19 data quality was estimated at 35.9% (95%CI=32.6%, 39.4%) while the rate of the data dimensions of timeliness, completeness, and accuracy were 50.2% (95%CI=46.7%, 53.8%), 50.6% (95%CI=47.1%, 54.2%), and 72.4% (95%CI=69.1%, 75.5%) respectively. It was found that the availability of a functional data validation team at the health facilities (AOR=18.3; 95%CI=1.62, 20.7; p= 0.019), training of data managers in COVID-19 data management (AOR=9.37; 95%CI=2.56, 34.3; p=0.001), and data managers with two-year professional training (certificate background) (AOR=3.42; 95%CI=1.95, 12.2; p=0.025) were independently associated with COVID-19 data quality.

**Conclusion:** The overall COVID-19 data quality in the Ahafo region was quite poor. Dimensionally, while the rate of data timeliness was high, that of data completeness, and accuracy were relatively low. The interaction of the independent correlates of COVID-19 data quality requires the healthcare system to identify stringent measures to strengthen the health information system to enhance planning and decision-making, especially during disease outbreaks.

## Introduction

Assessing the progress toward the Sustainable Development Goals (SDGs), World Health Organization (WHO) Triple Billion targets, and national or subnational health priorities requires a well-functional and proactive health information management system (HIMS) with high-quality data, especially in developing countries (1,2). Therefore, in ensuring public health efforts in combating pandemics, transparent data reporting that is of high quality and easily accessible is crucial for addressing outbreaks like coronavirus disease 2019 (COVID-19) (2–4). The WHO recommends four data quality dimensions including completeness, accuracy, consistency, and timeliness that need to be ensured at all levels of reporting (5). The repercussions of poor data quality in the health sector are significant, as they undermine efficient resource allocation and jeopardize national and international investments (2,6). Inaccurate data minimizes the effective monitoring of pandemics, global immunization initiatives and also increases the risk of vaccine-preventable disease outbreaks by failing to identify areas with low vaccination coverage (7,8). In the United States of America (USA), poor data quality is the third leading cause of death (9).

Globally, as data quality presents a significant concern, approximately 40% of countries lack evident adherence to quality data assurance processes which are mainly collected by national HIMS (2,5). For example, as of July 6, 2020, France had incomplete COVID-19 data on ages, while Armenia, Luxembourg, North Macedonia, Turkey, Serbia, and Bosnia-Herzegovina had no age data available. Similarly, as of May 1, 2020, the United States of America lacked a standardized system for COVID-19 data reporting, leading to significant discrepancies in data quality among different states and counties (8,10). Although there is no perfect health data due to missing values, bias, measurement, transcription, and human entry errors, most of these data problems chiefly occur in developing countries (11–14). Poor data quality in the health sector can be attributed to various factors, including inadequate training of data managers and healthcare staff, limited access to resources such as technology and reporting tools, and data fragmentation due to disparate systems (2,11,15,16). Data entry errors and lack of supervision and quality control also contribute to inaccuracies, alongside overwhelming workloads and time constraints that lead to rushed data entry (8,9,15). Furthermore, the absence of a dedicated data validation team, inconsistent reporting practices, and the lack of data governance and standardization further compromise data quality (7,17).

COVID-19 disease, caused by severe acute respiratory syndrome coronavirus-2 (SARS-CoV-2) was declared a pandemic and non-Public Health Emergency of International Concern in March 2020 and May 2023 respectively by WHO (3). The COVID-19 pandemic required continuous reporting formats via the Surveillance Outbreak Response Management and Analysis System (SORMAS) and/or HIMS across the globe (3,8). The Ghana Health Service under the Ministry of Health implemented the District Health Information Management System (DHIMS-2) software in 2008 to assist all districts, regions, and national managers in the analysis of routine health service data, planning, policy implementation, and also for enhancing health information reliability (6,18). In 2020, SORMAS, another web-based tool was also integrated into DHIMS-2 for reporting COVID-19 data. The DHIMS-2 utilizes three data dimensions (completeness, accuracy, and timeliness) in assessing data quality (18) which is employed in this study. Endriyas and colleagues re-echoed that completeness, accuracy, and timeliness are the most crucial indicators of data quality measurements (15). When these indicators are corrected, the other possible dimensions of data quality will automatically be addressed (15,19).

Ahafo Regional Health Directorate is one of the recently created directorates in Ghana which is confronted with resource constraints and inadequate technical human resources (20). The directorate was automatically hooked onto the DHIMS-2 and started recording high COVID-19 cases after the first case was registered in March 2020, the same month the country recorded its first incident (21). Data documentation and reporting of COVID-19 diseases then took center stage, although numerous studies have shown that data from Ghanaian health facilities are of poor quality in terms of accuracy and timeliness (18,22,23). This highlights the need to assess COVID-19 data quality using the three dimensions of data quality at different levels of the healthcare system. Such a study will not only furnish health managers in the Ahafo Region and across the 16 regions in Ghana with concrete ideas and knowledge on COVID-19 data quality challenges in the DHIMS-2 but will also recommend strategies to strengthen data quality for routine healthcare reporting and unforeseen disease outbreaks. Additionally, since the creation of the directorate in May 2019 (20), and the adoption of DHIMS-2 as a standard tool for reporting health data (21), no studies have been conducted in the Ahafo Region and Ghana at large to evaluate the quality of data on DHIMS-2, especially in the era of the COVID-19. In this regard, this study seeks to assess the determinants of COVID-19 data quality in the DHIMS-2 of the Ahafo Region in the middle belt of Ghana.

## Methods and materials

### Study setting

The study was conducted at Ahafo Region which was carved out of the southeastern part of the former Brong Ahafo Region of Ghana. It covers an estimated land area of 5,193 km^2^ and is surrounded by Ashanti, Bono, Brong Ahafo, and Western North Regions. This region lies within the tropical forest zone and is a key cocoa and timber-producing area. Two main reservoir systems could be distinguished in the region: the Bia and Tano rivers which also serve as the source of water supply to the region. There are six districts (Asunafo North, Asunafo South, Asutifi North, Asutifi South, Tano North, and Tano South) in the region. The region’s projected population is 576,526 as of 2022, with a growth rate of 2.3% from the 2021 population and housing census (24). There are 12 hospitals, 22 health centres, 16 clinics, 60 community health-based preventive services (CHPS), and three maternity homes located in the region. Out of these 120 health facilities, eight, 15, and 97 of them are owned by missionaries, private, and government respectively (20). Meanwhile, only 23 of these health facilities reported on COVID-19 cases in this region from 2020 to 2022 (21).

### Study design and sampling

This study employed a facility-based cross-sectional study design with a quantitative approach. We assessed and reviewed all 782 monthly reports of COVID-19 from March 2020 to December 2022 in all 23 health facilities. The monthly reports on COVID-19 were initiated in March 2020, however, the dataset was introduced into DHIMS-2 in July 2020. Non-COVID-19 monthly reports in the DHIMS-2 were excluded from this study. Moreover, all the data managers responsible for submitting monthly COVID-19 reports and having signed all the monthly reporting forms were interviewed.

### Data collection

The data collection period was from 6^th^ February to 30^th^ March 2023. Four trained research assistants were recruited to collect the data. A structured questionnaire was used to collect information on a computer-assisted interviewing device (android tablet or phone) through face-to-face interviews with data managers. Information on data managers’ characteristics (cadre, work experience, educational level, and training on SORMAS and COVID-19 data management), and health facility characteristics (availability of reporting tools, data validation team, registers, internet access, and supportive supervision) were collected. A comprehensive Excel sheet (collation sheet) was used to extract the data on completeness, accuracy, and timeliness from the registers, report forms, and DHIMS-2.

### DHIMS-2 and registers review

Three data collation sheets were used to collect the data on completeness, accuracy, and timeliness in all 23 health facilities over a retrospective 34-month starting from March 2020 to December 2022. These reviews were based on previous studies (5,6,11,15,23,25). The first collation sheet examined the completeness of COVID-19 data in the DHIMS-2 to see whether all data elements were filled. There are 102 data fields on the COVID-19 form to be completed. If one or more fields were left blank, that whole report for the month was deemed incomplete and therefore scored zero. On the other hand, if all the 102 data fields were filled the report for that month was deemed complete and therefore scored one. The second collation sheet was used to assess data accuracy by comparing COVID-19 data values from DHIMS-2, the COVID-19 monthly reporting form, and the COVID-19 register to determine the data accuracy. If the data values vary across the three data sources, the report for that month was deemed inaccurate and therefore scored zero. On the other hand, if the data values are the same across the three sources, the report for that month was deemed accurate and therefore given one score. The third collation sheet was used to extract the timeliness rate directly from DHIMS-2 for each month. Data entered into DHIMS-2 after the 15th of the ensuing month were deemed untimely. Finally, the data quality was determined when a monthly report scores three (thus, one score each for completeness, timeliness, and accuracy) (6). Quality and non-quality data were coded as “one” and “zero” respectively.

### Study variables

The dependent variable was COVID-19 data quality coded as “1 = data quality” and “0 = no data quality” (15). Based on earlier studies (11,15,17), the independent variables were made up of data managers’ information and facility-based characteristics as shown in Table 1.

### Data analysis

The data analysis was conducted using the statistical software, STATA version 17.0 (Stata Corp LLC, College Station, TX, USA). Descriptive statistics such as the rates of data quality including completeness, accuracy, and timeliness were computed (18). Also, a trend analysis of the data quality rates of completeness, accuracy, and timeliness was performed. Data accuracy was determined when the monthly COVID-19 report met all three data measures of completeness, accuracy, and timeliness (15). Independent variables were grouped or categorized where necessary (11,15,17). The independent variables were multi-level nested by facility data managers and all health facilities. Spearman’s correlation coefficient was used to check for multicollinearity among the independent variables before logistic regression analyses. Mixed-effects logistic regression was then performed to identify the possible determinants of the COVID-19 data quality.

### Ethical declarations

Two ethical approvals were obtained from the Institutional Review Board, School of Tropical Medicine and Global Health, Nagasaki University-Japan (NU_TMGH_2022_229_1), and Ghana Health Service Ethics Review Committee (GHS-ERC) with approval number GHS-ERC 033/01/23. Additionally, written permissions were obtained from the Ahafo Regional and all District Health Directorates. Data managers were made to sign the written informed consent after explaining the content to them.

## Results

### Descriptive analysis of COVID-19 monthly data quality from 2020 to 2022

All the 782 COVID-19 monthly reports comprising 34 months from 23 health facilities were collected and reviewed. The overall rates of data completeness, accuracy, and timeliness of the monthly COVID-19 report were 50.6% (95%CI=47.1%, 54.2%), 50.2% (95%CI=46.7%, 53.8%), and 72.4% (95%CI=69.1%, 75.5%) respectively. Meanwhile, the overall data quality was estimated at 35.9% (95%CI=32.6%, 39.4%) (Table 2).

Furthermore, the trend analysis of data completeness, accuracy, and timeliness in the region is illustrated in Figure 1. Data completeness of the COVID-19 report was from 30% in March 2020 which gradually increased to 70% in May and June 2022, before declining steadily to 57% in December 2022. Furthermore, the rate of data accuracy across the 34 months was generally low, starting from 18% in March 2020, with several rises and falls in between, before reaching 50% in December 2022. The records review at the various facilities revealed that COVID-19 reporting was introduced into the DHIMS-2 in July 2020 which accounted for the zero-timeliness rate from March to June 2020. Compared with data completeness and accuracy, the rate of data timeliness was generally high. The trend of data timeliness started at 73% reaching its lowest rate (43%) in December 2020 and its highest rate of 100% in August and September 2022, before declining to 83% in December 2022.

**Figure 1:**
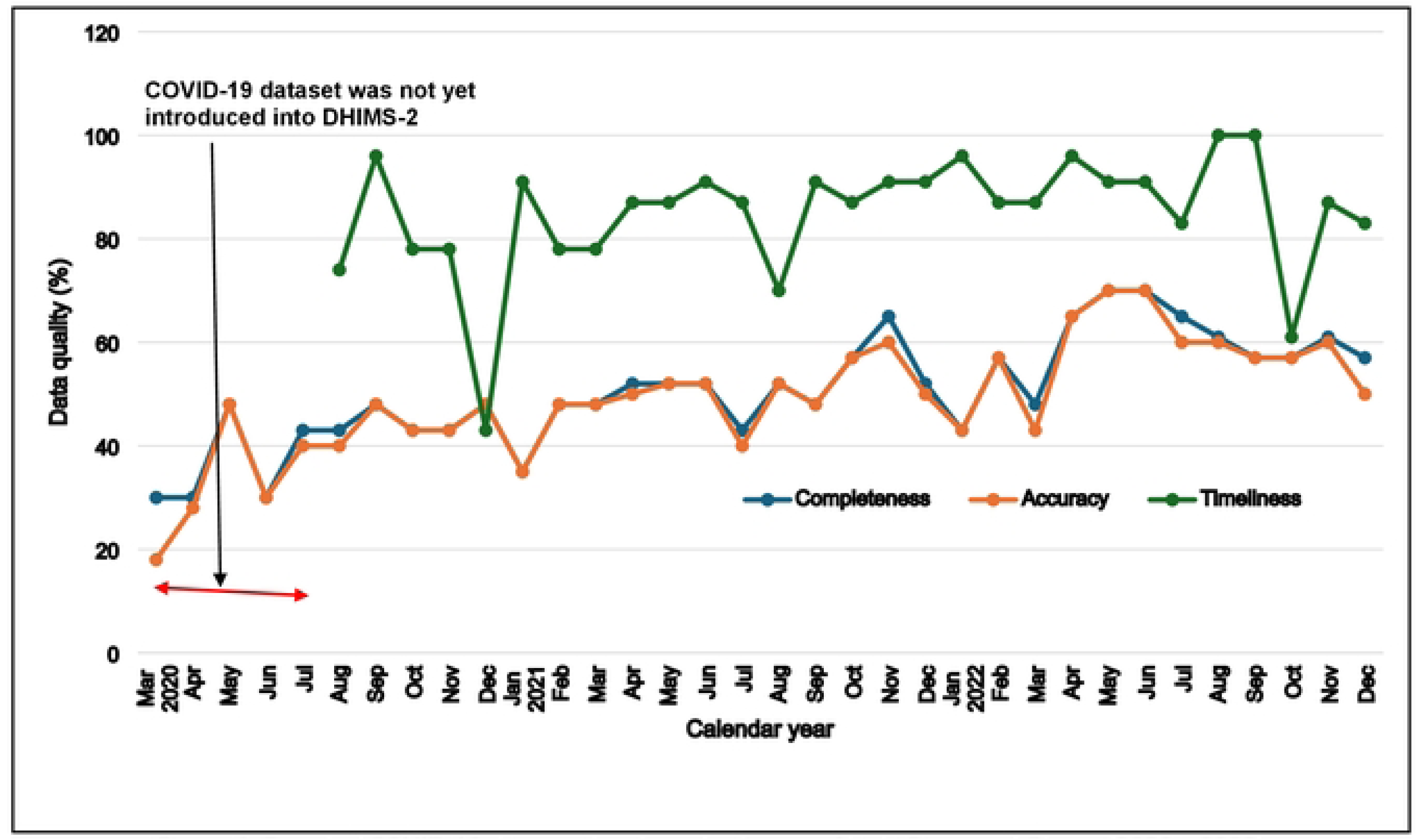
Trend of COVID-19 data quality from facilities in Ahafo Region of Ghana (2020 - 2022)

### Descriptive analysis of health facilities and data managers’ characteristics on COVID-19 reports from 2020 to 2022

The characteristics of health facilities and data managers are shown in Table 3. From the review of the 782 COVID-19 reports at the health facilities in the region, a large proportion of the reports were from health centres (56.5%) and government-owned health facilities (86.9%). The study revealed that most of the reports were produced and submitted by data managers who were males (73.9%), were health information officers (47.8%), and attained a diploma level of education (34.8%). Out of the total reports, 65.2% and 60.9% were submitted by data managers trained on COVID-19 data management and the SORMAS tool respectively. Moreover, of the total monthly reports submitted from 2020 to 2022, most of the reporting health facilities have data validation teams (95.7%), computers (87.0%), reporting registers (82.6%), internet access (78.3%), and inadequate reporting forms (60.9%) to support the activities of the data managers.

The mean(±SD) working experience among the data managers was 7.0(±5.7) years. Although the recommended number of HIOs per health facility in Ghana is two, the mean HIOs per facility was 1.53(±1.9) which could have affected the data quality in the region. Whilst the mean(±SD) outpatient department (OPD) attendance was 1,798(±197) per facility, that of suspected COVID-19 cases was 3.54(±1.9).

### Mixed effect analysis on determinants of COVID-19 data quality

Mixed-effect logistic regression was performed to identify some determinants of COVID-19 data quality in the Ahafo region of Ghana (Table 3). Five plausible variables showed statistical significance in the univariate mixed-effect analysis. There was no multicollinearity issue among the five variables after multicollinearity testing, so all were forwarded into the multivariate mixed-effect logistic regression model. After analysis, three variables including training on COVID-19 data management, data managers with certificate background, and availability of data validation teams were statistically significant in the final model.

The results revealed that the odds of reporting quality COVID-19 data into DHIMS-2 every month is 18.3 times higher when there is an available data validation team at the health facilities as compared to those without a validation team (AOR=18.3; 95%CI=1.62, 20.7; p= 0.019). In addition, data managers trained in COVID-19 data management were more likely to report quality data as compared to untrained ones (AOR=9.37; 95%CI=2.56, 34.3; p=0.001). Data managers having a certificate background in the profession were 3.42 times more likely to report higher data quality into DHIMS-2 as compared to those with a bachelor’s degree (AOR=3.42; 95%CI=1.95, 12.2; p=0.025).

## Discussion

The provision of safe and dependable healthcare relies on the availability of high-quality data (4). In addition, the monitoring of health performance is heavily dependent on quality data obtained from health facilities (2). This study aimed to assess the estimates and determinants of COVID-19 data quality in DHIMS-2 in the Ahafo Region of Ghana, by reviewing all COVID-19 monthly reports from March 2020 to December 2022.

Most of the COVID-19 reports (87%) were submitted by government-owned health facilities with few reports from mission and private facilities. This was expected as the majority of mission and private facilities in the region could not conduct COVID-19 tests during the early stages of the COVID-19 pandemic, therefore all suspected cases were sent to government-owned facilities for testing and possible management. Although there are several studies about COVID-19 in general (8,11), no or very scanty published studies on COVID-19 data quality are available which renders this study unique. Therefore most of these data quality estimates in this study were compared with other health indicators like maternal and child health as well as expanded program on immunization.

We found the completeness, accuracy, and timeliness of COVID-19 data reports in the region to be 50.6%, 50.2%, and 72.4% respectively with an overall data quality of 35.9%. Ouedraogo and colleagues reported data completeness and timeliness of 34% and 32% on selected MCH indicators respectively in Kersa City, Ethiopia which is less than our study results (26). Additionally, another study in sub-Saharan Africa reported the data accuracy rate of confirmed malaria as 46.4%, and that of antenatal care fourth visit as 46.6% (15) which are all lesser as compared to the results of the current study. The plausible reason for these indifferences could be attributed to greater political commitment to COVID-19 as compared to the MCH indicators which are regarded as routine data in healthcare delivery. Notwithstanding, the overall data quality of MCH indicators in Central Ghana was estimated at 93% (23) which is higher than our study estimate of 35.9%. Despite the political commitment to COVID-19 management, the disparity may be due to the higher number of monthly data reports and the novelty nature of COVID-19. More COVID-19 data was assessed and reported over a period of three years which could contribute to reducing COVID-19 data quality compared to MCH data reports assessed over 12 months to determine its quality in the previous studies.

Health facilities having functional data validation teams were more likely to report quality COVID-19 data (complete, accurate, and timely data) onto DHIMS-2. Coherent to studies in Tanzania, health facilities with data validation teams remain a significant determinant of data quality (27). Among the Ghanaian health facilities, data validation teams are mandated to meet within the first week of every month to review and validate the monthly report of the previous month before it is submitted onto the web-based DHIMS-2 platform. The activities of the data validation team do not only reduce data errors but also enhance the quality of data produced in the health facilities for planning and decision-making (6).

Training on COVID-19 data management was a significant factor for data quality in the Ahafo region which is parallel with the findings of other studies (7,17,28). Data managers should have adequate knowledge about data standards, best practices, and validation techniques to maintain data completeness, accuracy, and timeliness. Understanding data governance, compliance requirements, and ethical considerations is crucial for data privacy and security (2,6). Therefore, trained data managers are more likely to implement quality assurance and control measures to generate quality data at all levels and at all times (5,11). In this study, more than half of the facility data managers (65.2%) received training on COVID-19 data management which requires additional effort to build the capacity of the untrained ones in the Ahafo region. We therefore recommend on-the-job training for all facility staff responsible for data management as capacity development is key to quality data outcomes (2,28).

Data managers possessing a certificate background in the profession were about four times more likely to report quality data into the DHIMS-2. Though this is an unexpected finding, however, the COVID-19 data in most lower-level health facilities such as health centres and CHPS compounds in the Ahafo region were managed and formally reported by enrolled nurses and community health nurses (20). These nurses are mostly certificate holders with two years of school training. Furthermore, these nurses doubling as data managers are well-trained in COVID-19 data management and frequently receive supportive supervision from their senior colleagues and other data managers which makes them more superior in reporting quality data into the DHIMS-2 database.

## Limitations of the study

The study encountered some limitations. Our study assessed limited data quality dimensions. Thus, the study assessed three data dimensions (completeness, accuracy, and timeliness) out of the four and six data quality dimensions recommended by WHO (5) and the Data Management Association (DAMA – UK) International (25) respectively. Although COVID-19 was a pandemic that lasted for some time, this omission might result in overlooking other crucial dimensions that could impact the overall data quality.

We assessed 20% (23/120) of health facilities in the region which might affect the generalizability of the findings. The results might not be fully representative of all health facilities in the Ahafo Region of Ghana, which could limit its applicability to other contexts.

Lastly, despite adequate training of data collectors, there could be human or review errors (reporting bias) that might have occurred during the data collection process, which could introduce some inaccuracies in the study results.

## Conclusion

The overall COVID-19 data quality from 2020 to 2022 in the Ahafo region was quite poor. While the rate of data timeliness was high, that of data completeness, and accuracy were relatively low. The quality of COVID-19 data for the period was determined by the availability of a functional data validation team at the health facilities, training of data managers in COVID-19 data management, and data managers with two-year professional training (certificate background). The rate and interaction of these independent correlates of COVID-19 data quality require the healthcare system to identify stringent measures to focus on HIMS for planning and decision-making, especially during disease outbreaks and/or pandemics. Continuous training programs, workshops, and capacity-building initiatives for data managers with the necessary expertise in data collection, validation techniques, data entry protocols, and familiarity with reporting systems such as DHIMS-2 should be promoted. By investing in their professional growth, the GHS can empower data managers to adhere to data quality standards and ensure accurate and reliable data. Essential data tools such as reporting forms and registers should also be readily available at all times across health facilities. A dedicated data validation team should be established across all health facilities, and districts health directorates to systematically review and validate data before it is entered into the reporting database system. Providing specialized training to team members on data validation techniques and quality assurance procedures is crucial to ensure high-quality data during epidemics.

## Data Availability

The datasets collected, generated, or analyzed during this study have been attached as supplementary information.

## Acknowledgments

We appreciate The Project for Human Resource Development Scholarship - Japan International Cooperation Agency (JICA); Nagasaki University School of Tropical Medicine and Global Health Japan; and Ahafo Regional Health Directorate (Ghana Health Service) – Ghana, for their unwavering support of this study. The authors are also grateful to all data collectors, data managers, and volunteers who hugely contributed to this study.

## Abbreviations

COVID-19: Coronavirus disease 2019
DHIMS-2: District Health Information Management System version 2
HIMS: Health information management system
SARS-CoV-2: Severe acute respiratory syndrome coronavirus-2
SDGs: Sustainable Development Goals
SORMAS: Surveillance outbreak response management and analysis system
WHO: World Health Organization

## Consent For Publication

Not applicable

## Authors’ contributions

FGA, SAG, TA, and HA designed and conceptualized the study. FGA, SAG, SKBB, and HA supervised the data collection exercise which was carried out and supported by CA, SMB, and BBK. FGA, SAG, CA, SMB, BBK, SKBB, and HA performed the data analysis and interpretation. FGA, SAG, TA, and HA initially drafted the manuscript. All authors reviewed and approved the final manuscript.

## Authors’ acronyms

Felix Gumaayiri Aabebe (FGA); Silas Adjei-Gyamfi (SAG); Clotilda Asobuno (CA); Samuel Malogae Badieko (SMB); Blaise Bagyliku Kuubabongnaa (BBK); Samuel Kwabena Boakye-Boateng (SKBB); Tsunoneri Aoki (TA); Hirotsugu Aiga (HA)

## Supporting information

S1 Figure 1. Trend of COVID-19 data quality from facilities in Ahafo Region of Ghana (2020–2022)

S2 Table 1. Measurement and description of variables

S3 Table 2. Estimates of COVID-19 data quality from facilities in Ahafo Region of Ghana (2020–2022)

S4 Table 3. Frequency distribution and determinants of COVID-19 data quality from facilities in Ahafo Region of Ghana (2020–2022)

